# Transformer-based multiclass segmentation pipeline for basic kidney histology

**DOI:** 10.1101/2025.03.16.25324049

**Authors:** Junling He, Pieter A. Valkema, Jingmin Long, Jia Li, Sandrine Florquin, Maarten Naesens, Priyanka Koshy, Tri Quang Nguyen, Soufian Meziyerh, Aiko P J de Vries, Onno J de Boer, Verbeek Fons J, Zhan Xiong, Jesper Kers

**Affiliations:** Department of Pathology, LUMC, Leiden, Netherlands; Leiden Institute of Advanced Computer Science, Leiden University, Leiden, Netherlands; Department of Pathology, Amsterdam UMC, Amsterdam, Netherlands; Department of Nephrology and Transplantation, KU Leuven, Leuven, Belgium; Department of Microbiology, Immunology and Transplantation, KU Leuven, Leuven, Belgium; Department of Pathology, UMC Utrecht, Utrecht Netherlands; Division of Nephrology, Department of Medicine, LUMC, Leiden, Netherlands; Leiden Transplant Center, LUMC and Leiden University, Leiden, Netherlands; HIMS, University of Amsterdam, Amsterdam, Netherlands

**Keywords:** Deep learning, Kidney histology, Multiclass segmentation, Semantic segmentation, Transformer

## Abstract

Multiclass segmentation of microanatomy in kidney biopsies is an important and non-trivial task in computational renal pathology. In a multicenter study, we densely annotated basic anatomical objects (glomeruli, tubules, and vessels) in 261 regions of interest of 147 kidney biopsy WSIs sourced from the archives of hospitals in Amsterdam, Utrecht, and Leiden (Netherlands). And we trained multiple UNet- and Mask2Former-based models on WSI-level and patch-level splitting methods, and compared their performance across training strategies. Test performance was assessed on 24 annotated renal WSIs from Leuven (Belgium) with sensitivity analysis on the extent of fibrosis and inflammation.

At the patch-level, UNet-ResNet18 achieved comparable performances to M2F-Swin-B with average Intersection over Union of all classes (A-IoU, 0.84 vs 0.94), as well as per-class IoU. However, at the WSI-level, M2F-Swin-B significantly surpassed UNet-ResNet18 with large margins on A-IoU (0.84 vs 0.48), with similar observed in per-class IoU. Notably, M2F-Swin-B outperformed UNet-ResNet18 in scenarios characterized by a higher degree of fibrosis and inflammation (A-IoU, 0.76 vs 0.66). Furthermore, at the WSI-level, M2F-Swin-B achieved IoU score of arteries to 0.58, whereas UNet-ResNet18 only achieved 0.33. In this study, we found that the attention mechanism in Mask2Former enables visibly crisper and more uniform segmentation, particularly when data is inadequate. Mask2Former-based models outperform UNet-based models in challenging areas from inflamed and fibrotic renal biopsies.

## Introduction

Renal pathology is a subspecialty of general pathology focused on characterizing and diagnosing nephrological diseases. The kidney biopsy is the gold standard for diagnosing and staging kidney diseases. Absolute quantification of every lesion in the renal biopsy, independent of disease entity, is the holy grail of quantitative renal pathology. However, the evaluation of kidney biopsies predominantly depends on manual visual analysis, which inevitably introduces inter-observer variability[1]. Full manual quantification and annotation of abnormalities in repetitive structures like tubules is not feasible within the diagnostic time frame. Consequently, renal pathologists are compelled to score (i.e. estimate) tubular lesions using non-granular, semi-quantitative ranges like 25-50%. Nonetheless, there may be significant differences in prognosis between patients with 26% and 50% tubular injury. Additionally, each underlying disease has its lesion-scoring scheme, despite the general acceptance that kidneys respond to injury in stereotypical patterns that can co-occur within a single structure (e.g., crescents appear morphologically identical across different kidney diseases) [2].

With the rapid development of big data, the continuous emergence of new biotechnology, and the innovation of artificial intelligence (AI) algorithms, the field of nephrology has experienced significant progress and changes[3, 4]. AI-assisted renal pathology offers a promising solution for pathologists to address the tedious process of quantifying thousands of individual lesions. Since the basic micro-anatomy of the kidney does not change during the progression of any disease, a high-performing and robust multiclass segmentation network could be the first step toward building a community effort for quantitative renal pathology.

Most recent research about deep learning (DL) applications on renal histopathology can generally be categorized into two main areas: object detection[5–8] and segmentation[9–15]. As DL methods have advanced, architectures such as U-Net[14–16], DeepLab[13,17], SegNet[15], and Cascade Mask R-CNN[18] have been effectively employed to address challenges within both semantic and instance segmentation. While the DL applications for segmenting anatomical compartments of the kidney are increasing, this basic operation remains challenging due to two primary issues: 1) most models cannot achieve state-of-the-art performance because a lack of high-quality annotated data, especially in areas with significant kidney damage, fibrosis, and inflammation, and 2) there are currently no publicly available kidney segmentation DL models. Beyond the DL applications on segmenting anatomical compartments and scoring lesions in renal histology, recent studies have demonstrated the potential for high-throughput morphological analysis of kidney histology, termed Next Generation Morphometry, which aligns with high-throughput techniques like next-generation sequencing in preclinical experiments for various kidney diseases[19].

The most commonly used DL models for segmenting anatomical compartments of the kidney are convolutional neural networks (CNNs). In contrast, Transformers are newer architectures based on attention mechanisms[20]. Originally designed for natural language processing tasks, Transformers have since been adopted for various computer vision tasks, including segmentation. Unlike CNNs, which accumulate local features through a stack of convolutional layers, Transformers can directly capture long-range dependencies to form global features. In 2021, Cheng et al. developed a Transformer-based segmentation architecture called Mask2Former[21], which has become a powerful and popular option for segmentation models, outperforming conventional CNN-based architectures on various segmentation tasks.

This study primarily aims to create a broadly applicable open-source segmentation pipeline that is robust against the heterogeneity of clinical kidney biopsy presentations. This pipeline is intended to serve as a foundation for future research and drive innovation in the field of computational nephropathology. We test the performance of CNN- and Transformer-based segmentation networks on a multicenter dataset, focusing on renal WSIs with varying degrees of interstitial inflammation and fibrosis. We found that Mask2Former, especially when equipped with a Swin-B Transformer encoder, consistently outperformed the CNN-based UNet segmentation models in cases of inflammation and fibrosis, including smaller, hard-to-segment objects like arterioles. Lastly, the source code and the model weights are available online https://github.com/amspath after request.

## Materials and Methods

### Sample selection

A multicenter retrospective training set was created comprising Jones-silver stained kidney biopsy slides from 147 patients from the Departments of Pathology of the Amsterdam UMC (AUMC), UMC Utrecht (UMCU) and Leiden UMC (LUMC). All the slides were anonymized by a pathology staff member before further analysis. Kidney WSIs from both transplant and native kidney biopsies were digitized with a Philips UltraFast scanner (AUMC, LUMC) or Hamamatsu XR scanner (UMCU) at a resolution of ∼0.25 µm/pixel. One to five representative regions of interest (ROIs) on a single WSI were chosen for dense manual annotations, and a total of 261 ROIs were acquired for further annotations (**Figure 1A**). Instance object annotation details can be found in the supplementary materials.

**Figure 1.**
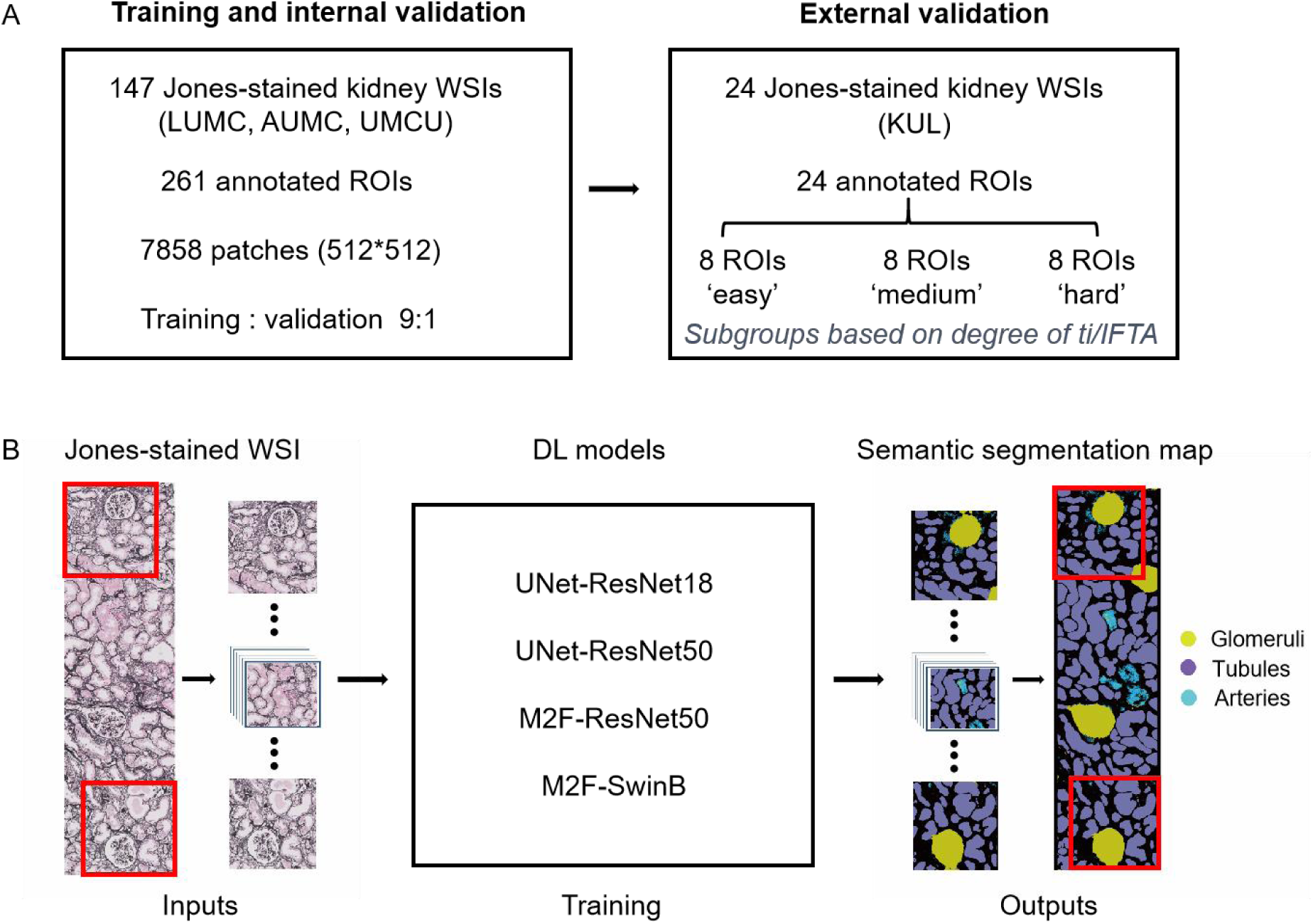
Study design. **A.** The details of the training, internal validation and external validation datasets used in the current study. **B.** The workflow of the current study. CNN-based (UNet-ResNet18, UNet-ResNet50) and Transformer-based (M2F-ResNet50, M2F-SwinB) architectures were trained to segment the kidney anatomical compartments on Jones-stained renal WSIs. M2F: Mask2Former.

An independent external test set was created with an anonymized collection of a total of 24 digital kidney biopsy cases from the Department of Pathology of KU Leuven (KUL), scanned with a Philips UltraFast scanner at a resolution of 0.25 µm/pixel. All external test cases were subgrouped according to the degree of interstitial inflammation and fibrosis according to the Banff 2022[22] ti- and IFTA scores according to the following definitions: ti/IFTA ≤25% were assigned as “easy biopsy” group (n = 8), ti/IFTA 26-50% were assigned as “medium biopsy” group (n = 8) and ti/IFTA >50% were assigned as “hard biopsy” group (n = 8). A representative ROI according to the pathology report from each WSI was chosen by a senior nephropathologist (**Figure 1A**).

This study was approved by the Research Ethics Committee of different hospitals (KUL approval number: S64006, LUMC approval number: W2020.031, AUMC approval number:19.260, and UMCU approval number:19.482).

### Instance object annotations

A total of four classes were densely annotated in each ROI until all pixels were assigned to a class: background, glomeruli, tubules, and arteries (inter-, intralobular arteries and arterioles altogether). The interstitium class (which includes interstitium/stroma and peritubular capillaries) was defined as the residual pixels after subtraction of all four positive classes (background, glomeruli, tubules, and arteries). This ensured that all pixels received a class label. To prevent the predicted tubules from merging due to close proximity, particularly in the relatively normal kidney biopsies, we introduced an auxiliary boundary class that was generated on the fly during training. Each boundary was eroded along the corresponding compartment mask with 4 pixels by a morphological erosion operation. All cases were annotated with ASAP version 1.9 (https://computationalpathologygroup.github.io/ASAP/) and our in-house custom WSI reader Slidescape, which can be freely downloaded at https://github.com/amspath/slidescape. Images were annotated by trained physicians who are experienced with deep learning pipelines and all annotations were validated by a nephropathologist with experience in computer vision algorithm training procedures. Annotations of difficult structures were discussed as a group.

### Pre-processing of WSIs for Training

A resolution of 0.5 µm/pixel was chosen to extract regions of interest (ROIs) from the annotated WSIs. Patches with fixed dimensions of 512×512 pixels were extracted from the ROIs using a sliding window stride of 256 pixels. For the edges of the ROIs, zero padding was added to round their sizes up to a multiple of the patch size and stride. If the patch images were smaller than 512×512 pixels, zero padding was used to extend them to the fixed dimensions. A total of 7,858 patch images were generated for the training dataset. During training, the padding regions were ignored. All patch images were randomly split into a training set (90%) and an internal validation set (10%). This split was performed at both the patch level and the WSI level (case level). The patch-level split may introduce information leakage from the training procedure to the validation procedure, potentially leading to more stable training. In contrast, the WSI-level split creates a larger domain shift from training to validation, which might increase regularization and potentially improve out-of-domain generalization. These patch- and WSI-level train-validation split settings were directly compared using the KUL external test set, which was always left out of the training and validation processes.

### Semantic segmentation model training

Two segmentation architectures were adopted in the current study. For CNN-based UNet network, we trained and tested the relatively shallow ResNet18 and the deeper ResNet50 encoder. For Transformer-based Mask2Former, we trained and tested the CNN-based ResNet50 encoder and the Transformer-based Swin-B encoder. The code was forked from the original UNet (http://lmb.informatik.uni-freiburg.de/people/ronneber/u-net) and Mask2Former implementations (https://github.com/facebookresearch/Mask2Former), and we conducted minimal adjustments to the architectures for the current application. All model training schedules were initialized with ImageNet-pretrained encoder weights. The default UNet decoder and Transformer decoder, default ADAMW optimizer, and default “WarmupPolyLR” learning rate scheduler were used. The UNet-ResNet models had a batch size of 16 and were trained for 50 epochs until convergence. Mask2Former models were trained with a batch size of 10 for 64 epochs until convergence. The Intersection over Union (IoU, i.e. Jaccard index) and pixel-level accuracy were used as an outcome metric, as the routine evaluation used by Mask2Former. A single NVIDIA RTX 4090 GPU was used for training all DL models. And the Post-processing and generation of new instance segmentation predictions can be found in the Supplementary materials.

### Post-processing and generation of new instance segmentation predictions

To reconstruct ROI-level results from patch predictions, the predicted patches were generated with an overlap of 0.5 patch stride. Patch predictions were subsequently stitched by a weighted average of overlapping areas, with weights from the 2-dimensional Hann window function. The peaks near the center of the patch were attenuated to 0 at the edges. Pixel-level class prediction were-rescaled using the *argmax()* function. Detectron2’s built-in visualizer code was used for visualizations of the predictions. In order to reuse the slide-level segmentation masks for further downstream annotation in Slidescape (e.g. glomerulosclerosis lesions), we propose a process to obtain new ground-truth annotations from the predicted masks and save them in XML format. The process consists of the following functions: (1) For predicted masks of each class, OpenCV *findContours()* is used to extract object boundaries; (2) *approxPolyDP()* with epsilon=1.0 is further utilized to approximate precise contours; (3) Then, inner contours are merged with outer contours to form a single object; (4) Lastly, objects with areas < 300 square pixels are filtered out. The auxiliary boundary class is ignored in the generation process.

## Results

### Cohorts and semantic segmentation models

The details of the training cohort, internal validation cohort and the external validation cohort are visualized in **Figure 1A**. As shown in Figure 1B, we selected CNN-based and Transformer-based models for a back-to-back comparison of segmentation to robustness against nephropathology heterogeneity. UNet-ResNet18, UNet-ResNet50, M2F-ResNet50 and M2F-SwinB were trained to segment kidney anatomical compartments, including background, glomeruli, tubules, arteries (inter-, intralobular arteries and arterioles altogether), and interstitium as a residual class. The baseline characteristics of the cohorts can be found in **Table 1**.

**Table 1.** Baseline Banff lesion scores of the cohorts.

|  | Total | AUMC | LUMC | UMCU | KUL | KUL | KUL |
| --- | --- | --- | --- | --- | --- | --- | --- |
| Difficulty group | - | - | - | - | Easy | Medium | Hard |
| Included in | Train + Val | Train + Val | Train + Val | Train + Val | Test | Test | Test |
| N (WSIs) | 147 | 49 | 58 | 40 | 8 | 8 | 8 |
| Glomeruli, median (IQR) | 20 (15-26) | 22 (18-32) | 19 (13-24) | 19 (13-23) | 24 (15-30) | 27 (22-36) | 23 (17-31) |
| Glomerulosclerosis, median % (IQR) | 31(0-29) | 11 (3-31) | 5 (0-18) | 15 (6-32) | 10 (4-13) | 23 (9-31) | 35 (15-41) |
| TMA, N (%) | 30 (20) | 11 (22) | 16 (28) | 3 (8) | 0 (0) | 0 (0) | 0 (0-0) |
| ATN/ATI, N (%) | 83 (56) | 28 (57) | 40 (69) | 15 (38) | 2 (25) | 4 (50) | 4 (50) |
| g-score, median (IQR) | 0 (0-1) | 0 (0-2) | 0 (0-1) | 0 (0-0) | 0 (0-0) | 0 (0-0) | 0 (0-0) |
| cg-score, median (IQR) | 0 (0-1) | 1 (0-2) | 0 (0-1) | 0 (0-0) | 0 (0-0) | 0 (0-0) | 0 (0-1) |
| mm-score, median (IQR) | 0 (0-1) | 0 (0-1) | 0 (0-0) | 0 (0-1) | 0 (0-0) | 1 (0-3) | 0 (0-0) |
| i-score, median (IQR) | 1 (0-2) | 0 (0-1) | 1 (0-2) | 1 (0-1) | 0 (0-0) | 0 (0-1) | 0 (0-2) |
| t-score, median (IQR) | 0 (0-2) | 0 (0-1) | 1 (0-2) | 1 (0-2) | 1 (0-1) | 1 (0-1) | 1 (1-2) |
| ti-score, median (IQR) | 2 (1-3) | 2 (1-3) | 2 (1-3) | 2 (1-3) | 1 (0-1) | 2 (1-3) | 2 (2-2) |
| IFTA-score, median (IQR) | 1 (1-2) | 1 (1-3) | 1 (0-2) | 2 (1-3) | 1 (1-1) | 2 (2-2) | 2 (1-2) |
| i-IFTA-score, median (IQR) | 2 (1-3) | 3 (1-3) | 2 (0-3) | 2 (1-2) | 1 (0-1) | 3 (2-3) | 2 (2-2) |
| t-IFTA-score, median (IQR) | 1 (0-1) | 1 (0-2) | 1 (0-1) | 1 (0-1) | 0 (0-0) | 2 (1-2) | 1 (1-1) |
| ptc-score, median (IQR) | 0 (0-0) | 0 (0-0) | 0 (0-1) | 0 (0-1) | 0 (0-0) | 0 (0-0) | 0 (0-0) |
| v-score, median (IQR) | 0 (0-0) | 0 (0-0) | 0 (0-1) | 0 (0-0) | 0 (0-0) | 0 (0-0) | 0 (0-0) |
| cv-score, median (IQR) | 1 (0-2) | 1 (0-2) | 1 (0-2) | 1 (1-2) | 1 (0-1) | 2 (1-2) | 1 (0-1) |
| ah-score, median (IQR) | 1 (0-2) | 1 (0-1) | 1 (0-2) | 1 (1-2) | 1 (0-1) | 2 (2-3) | 2 (1-2) |

WSIs = whole slide images; IQR = interquartile range; TMA = thrombotic microangiopathy; ATN/ATI = acute tubular necrosis/acute tubular injury.

### Mask2former-based networks outperform UNet-based networks on the internal validation dataset and external test dataset

UNet-based architectures are the *de facto* standard models in the vast majority of biomedical computer vision segmentation studies. We could observe that UNet-based architectures, a more shallow ResNet18 encoder outperforms a deeper ResNet50 encoder across all anatomical compartments when training and internal validation data are split at the patch-level (i.e. patches from the same WSI were shared between training and validation data): IoU 0.84 and 0.72, respectively (**Figure 2A**). Splitting the training and validation data at the WSI-level however resulted in a decline in IoU across all anatomical compartments with a slightly higher IoU for the UNet-ResNet18 models (IoU 0.48) over the UNet-ResNet50 model (IoU 0.41), albeit marginal, and still the IoU for individual compartments would favor the simpler ResNet18 encoder (**Figure 2B**).

**Figure 2.**
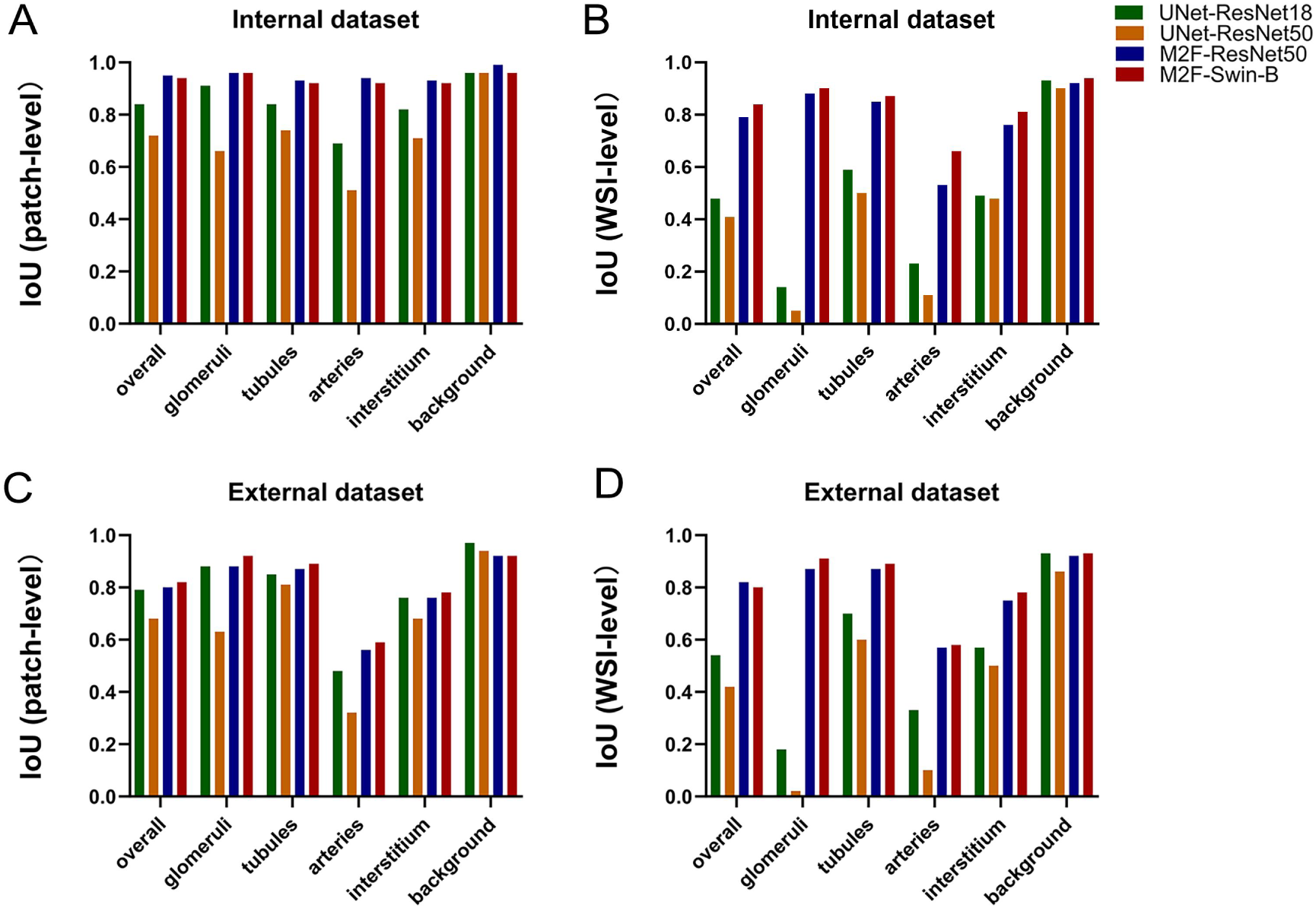
The performance of different DL models on the internal validation dataset with patch-level and WSI-level split. **A-B.** Overall IoU and IoU across all compartments of four models (UNet-ResNet18, UNet-ResNet50, M2F-ResNet50 and M2F-SwinB) in the internal dataset with the patch-level split (A) and WSI-level split (B). **C-D.** Overall IoU and IoU across all compartments of four models in the external dataset with the patch-level split (C) and WSI-level split (D). IoU: Intersection of Union; M2F:Mask2Former.

On the other hand, at the patch level, both Mask2Former models outperform the UNet models by a margin, which includes Mask2Former models with the CNN-based ResNet50 (IoU 0.95) encoder and SwinB encoder ((IoU 0.94) backbone (**Figure 2A**). This trend was observed across all histological compartments. Even though the performance in terms of IoU showed a tremendous drop for the UNet models when splitting at the WSI level, the IoU remained high for all histological compartments, with a slight preference for the fully Transformer-based M2F-SwinB model over the CNN-Transformer hybrid M2F-ResNet50 model: AC-IoU of 0.84 versus 0.79, respectively (**Figure 2B**). Of particular interest is the higher performance of the former model on the difficult-to-segment arteries compartment, which varies in size and has a relatively low pixel-label prevalence compared to the other classes: IoU 0.66 in M2F-SwinB model versus 0.53 in M2F-ResNet50 model, respectively (**Figure 2B**).

Furthermore, we observed ResNet18 encoder outperforms ResNet50 encoder across all histological compartments in the external test dataset, which is split at the patch level, with IoUs of 0.79 and 0.68, respectively (**Figure 2C**). However, splitting the test dataset at the WSI level resulted in a decline in IoU across all compartments, which is in line with the founding in the internal validation dataset (**Figure 2D**). Compared with WSI-level split, patch-level split training schedules in the internal dataset and external dataset has better stability, suggesting patch-level split introduces some leakage of information from the training procedure to the validation procedure. Nonetheless, the WSI-level split creates a larger domain shift from training to validation dataset, which might be the main reason for a decline in AC-IoU and IoU across all compartments.

### Mask2Former-based models outperform UNet-based models in challenging areas from inflamed and fibrotic renal biopsies

The external dataset was categorized according to the degree of ti-scores and IFTA-scores. **Figure 3** illustrates that the Mask2Former models outperformed the UNet models in terms of IoU across these subgroups. Interestingly, in this multiclass segmentation task, glomerular segmentation appeared to be difficult for the UNet models when splitting training and validation datasets at the WSI-level (especially for the model with the larger ResNet50 encoder, **Figure 3H**), which suggests a strong inductive bias for UNet-based renal pathology models due to between- and within-case pixel-label imbalance for this class.

**Figure 3.**
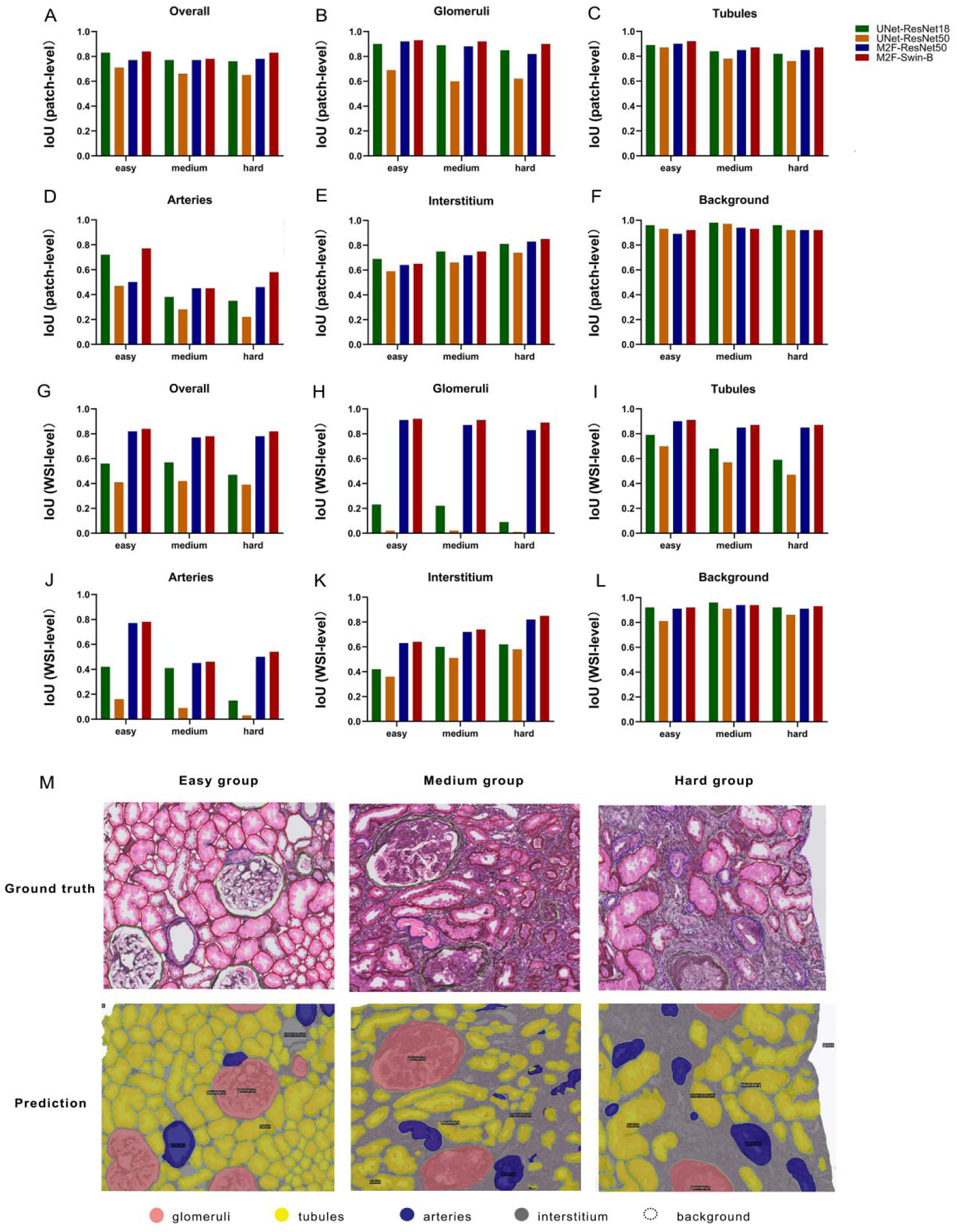
The performance of different models on the subgrouped external dataset. **A-F**. IoU of overall class, glomeruli, tubules, arteries, interstitium, and background in the subgrouped external dataset (easy group, medium group, hard group) for different models with the patch-level split. **G-L**. IoU of overall class, glomeruli, tubules, arteries, interstitium, and background in the subgrouped external dataset (easy group, medium group, hard group) for different models with the WSI-level split. **M**. The representative images of Jones-stained patches with ground truth annotations and the prediction masks generated by M2F-Swin-B in the subgrouped dataset. IoU: Intersection of Union; M2F: Mask2Former.

The UNet models faced particular challenges in identifying arteries in biopsies with a higher degree of inflammation and fibrosis, both for models trained in patch-level (**Figure 3D**) and WSI-level train-validation splits (**Figure 3J**). Similarly, the performance for the artery class on the external validation set in terms of IoU also declines for the Mask2Former segmentation models in both patch- and WSI-level train-validation splitting when the degree of ti-scores and IFTA-scores increased, albeit to less extent compared to the UNet models (IoU 0.54 and 0.50 for M2F-SwinB and M2F-ResNet50 vs IoU 0.15 and 0.03 for UNet-ResNet18 and UNet-ResNet50 in the hard group, respectively). Representative ground truth annotations and model prediction masks for the easy, medium and hard groups are visualized in **Figure 3M**.

### Extension of the basic anatomical segmentation model with pathological lesions

Next, we performed an extension to show a proof-of-concept how researchers can build their own models on top of the basic anatomical segmentation model. Since two Mask2Former models showed superior performance in segmenting the basic anatomical compartments, we selected these models for retraining the multiclass segmentation model with tubular atrophy and glomerulosclerosis included. **Figure 4A** and **4B** depict the performance of the two Mask2Former models on the 7-class semantic segmentation task, indicating a similar trend of the improved performance of the fully Transformer-based M2F-SwinB model (AC-IoU of 0.76) over the hybrid M2F-ResNet50 model (AC-IoU of 0.73) in the internal dataset. Specifically, the M2F-SwinB model achieved IoU of 0.75 for glomerulosclerosis and 0.55 for tubular atrophy in the internal dataset, and IoU of 0.84 for glomerulosclerosis and 0.38 for tubular atrophy in the external dataset. Examples of regions with tubular atrophy predicted by the M2F-SwinB model are illustrated in **Figure 4C**.

**Figure 4.**
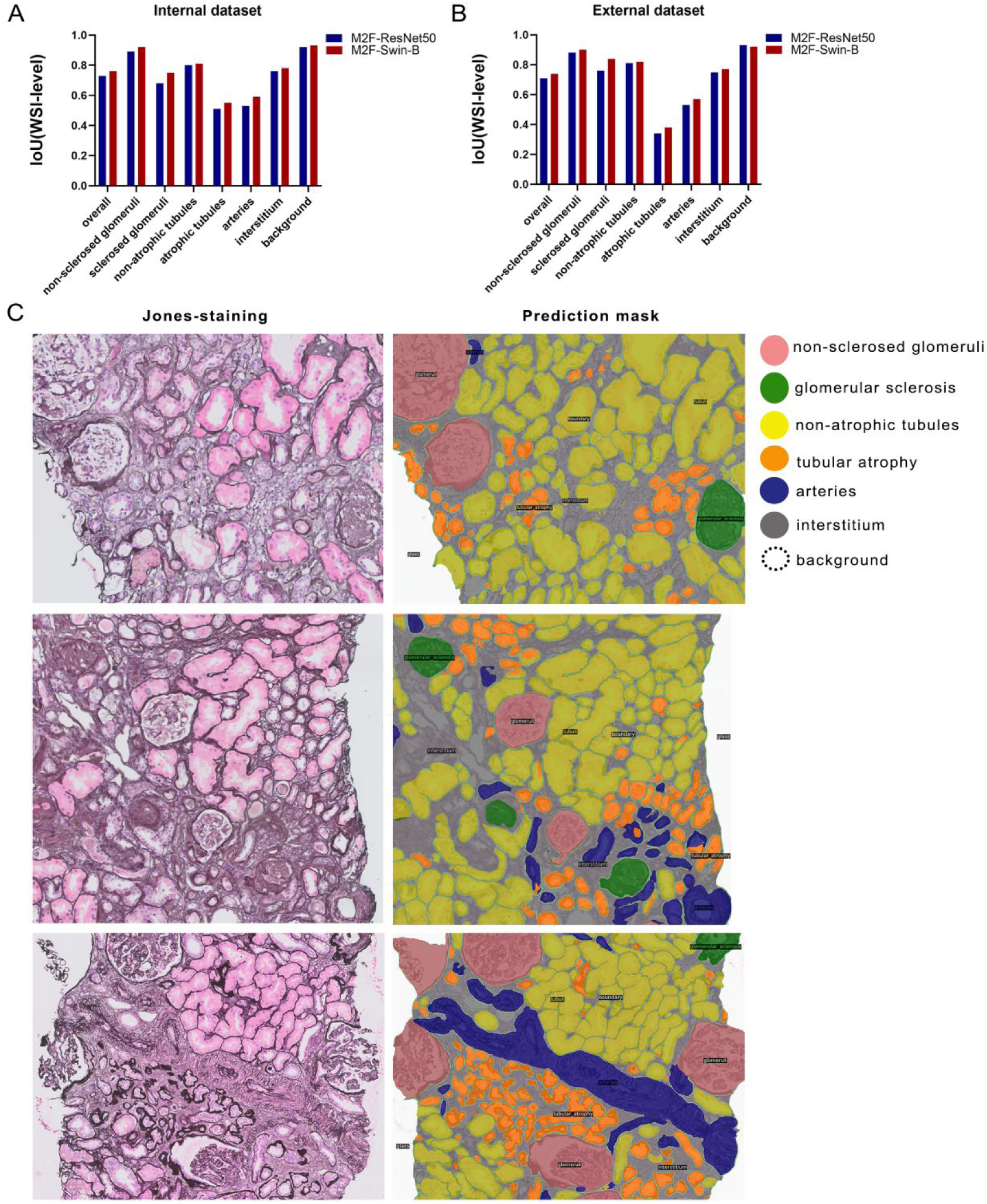
The performance of Mask2Former on segmenting the normal and abnormal kidney anatomical compartments. **A-B**. IoU of overall class, non-sclerosed glomeruli, sclerosed glomeruli, non-atrophic tubules, atrophic tubules, arteries, interstitium, background in two different Mask2Former models in the internal dataset (A) and external dataset (B) with the WSI-level split. **C**. The representative images of Jones-stained patch images and the prediction mask images generated by M2F-Swin-B model. IoU: Intersection of Union; M2F: Mask2Former.

### Histological scores that correlate with low segmentation accuracy

To better understand which histological factors negatively impact segmentation performance for the M2F-SwinB model, we performed linear regression analysis on (1 - IoU) calculated for each of the segmented compartments (macro-level, glomeruli, tubules, vessels, interstitium) on each of the 24 test cases (Table 2). On the macro-level, only the presence of acute tubular injury correlated to a lower IoU (beta = 0.09, SE = 0.001, p = 0.002). A lower glomerular segmentation IoU correlated to a lower number of glomeruli in the biopsy (beta = −0.006, SE = 0.002, p = 0.01). A lower tubular segmentation IoU correlated to a higher percentage of sclerosed glomeruli (beta = 0.003, SE = 0.001, p = 0.01), a higher degree of endothelialitis (beta = 0.21, SE = 0.07, p = 0.008) and to a higher degree of vascular intima thickening (beta = 0.04, SE = 0.02, p = 0.07). A lower vascular segmentation IoU correlated to the presence of acute tubular injury (beta = 0.27, SE = 0.11, p = 0.02) and a higher degree of arteriolar hyalinosis (beta = 0.10, SE = 0.05, p = 0.05). Lastly, a lower interstitium segmentation IoU correlated with a lower percentage of sclerosed glomeruli (beta = −0.003, SE = 0.002, p = 0.07), a lower degree of interstitial inflammation (beta = −0.06, SE = 0.02, p = 0.03), a lower degree of tubulitis (beta = −0.05, SE = 0.03, p = 0.07), a lower degree of total inflammation (beta = −0.06, SE = 0.02, p = 0.007), a lower degree of tubulitis in areas of IFTA (beta = −0.06, SE = 0.03, p = 0.05) and a lower degree of arteriolar hyalinosis (beta = −0.04, SE = 0.02, p = 0.07). Only the presence of acute tubular injury correlated to a lower IoU across all 4 compartments (**Table 2**).

**Table 2.** Histological factors contributing to a lower M2F-SwinB segmentation performance.

| Histological parameter | Macro<br>p-value | Glomeruli<br>p-value | Tubules<br>p-value | Vessels<br>p-value | Interstitium<br>p-value |
| --- | --- | --- | --- | --- | --- |
| Number of glomeruli | 0.94 (+) | **0.01** (-) | 0.17 (+) | 0.93 (-) | 0.10 (+) |
| Percentage of sclerosed glomeruli | 0.73 (+) | 0.53 (-) | **0.01** (+) | 0.25 (+) | **0.07** (-) |
| Acute tubular injury | **0.002** (+) | 0.12 (+) | 0.36 (+) | **0.02** (+) | 0.51 (+) |
| Glomerulitis (g-score) | 0.23 (-) | 0.48 (-) | 0.77 (-) | 0.21 (-) | 0.64 (+) |
| Transplant glomerulopathy (cg-score) | 0.57 (-) | 0.82 (+) | 0.66 (-) | 0.50 (-) | 0.52 (-) |
| Mesangial proliferation (mm-score) | 0.94 (-) | 0.40 (-) | 0.67 (-) | 0.68 (+) | 0.50 (-) |
| Interstitial inflammation (i-score) | 0.99 (+) | 0.25 (+) | 0.78 (-) | 0.35 (+) | **0.03** (-) |
| Tubulitis (t-score) | 0.17 (-) | 0.92 (+) | 0.88 (+) | 0.58 (-) | **0.07** (-) |
| Total inflammation (ti-score) | 0.84 (-) | 0.60 (+) | 0.18 (+) | 0.59 (+) | **0.007** (-) |
| Interstitial fibrosis/tubular atrophy<br>(IFTA-score) | 0.16 (+) | 0.76 (+) | 0.27 (+) | 0.20 (+) | 0.49 (-) |
| Interstitial inflammation in areas of IFTA<br>(i-IFTA-score) | 0.91 (+) | 0.32 (-) | 0.51 (+) | 0.27 (+) | 0.28 (-) |
| Tubulitis in areas if IFTA (t-IFTA-score) | 0.27 (-) | 0.33 (-) | 0.82 (-) | 0.93 (+) | **0.05 (-)** |
| Peritubular capillaritis (ptc-score) | 0.32 (+) | 0.99 (+) | 0.51 (-) | 0.70 (+) | 0.24 (+) |
| Endothelialitis (v-score) | 0.35 (+) | 0.71 (-) | **0.008 (+)** | 0.61 (-) | 0.35 (+) |
| Vascular intima thickening (cv-score) | 0.28 (+) | 0.45 (+) | **0.07 (+)** | 0.80 (+) | 0.77 (-) |
| Arteriolar hyalinosis (ah-score) | 0.36 (+) | 0.96 (-) | 0.19 (+) | **0.05 (+)** | **0.07 (-)** |

P-values in bold represent correlations trends with $P < 0.10$ . (+) indicates a positive association and (-) indicates a negative correlation.

## Discussion

As a companion to this article, we released 1) the code that was used to train the multiclass segmentation networks, 2) the model weights to either test the networks as is or use as initialization for fine-tuning on local data if needed, 3) code to stitch and recreate annotations from model prediction masks, and 4) the viewer software *Slidescape* that allows people to interact with the newly created annotations as objects and adjust them for further downstream applications.

Researchers have developed CNN-based frameworks to identify histopathological structures[13, 19, 23], segment glomerular cells[15, 24], quantify nonsclerotic and sclerotic glomeruli[14, 15, 18, 25–27], and assess IFTA grade[12, 28, 29]. Recently, Transformer architecture has also gradually demonstrated its robust capabilities in computer version[21]. In the current study, when ample data is available, patch-level splitting can be employed to assess the comprehensive performance of neural networks, as all patches from the entire set of WSIs can be accessed during the training. Conversely, the WSI-level splitting offers a stringent test of the generalization capabilities of neural networks under data-constrained conditions, as the patches for training and evaluation are compulsorily generated from different WSIs. After extensive evaluation in both internal and external datasets, we observed that the Transformer-based models significantly outperform CNN-based counterparts on WSI-level splitting, showing the strong domain transfer ability for large in-group data variation. In contrast, CNN-based models can achieve comparable performance on patch-level splitting with lower complexity. This contrastive approach not only illuminates the adaptability of the networks but also provides a benchmark for selecting DL models, contingent upon the availability of annotated datasets. In practical applications, the scarcity of data often poses a significant challenge, particularly within the specialized field of renal pathology. Consequently, the Transformer-based Mask2Former architecture, with its robust attention mechanism, could emerge as a more apt selection, especially in scenarios where annotated data is not abundant.

IFTA is a strong morphologic prognostic indicator of chronic kidney disease (CKD) progression and outcome, regardless of the etiology of the disease[30–38]. Given the myriad of tubules present and IFTA particularly occurs in a patchy fashion in the renal biopsy, it is impractical to manually enumerate the extent of atrophic tubules and the expanse of fibrosis and inflammation. Consequently, IFTA presents a continuous morphologic spectrum. Therefore, grading IFTA remains a manual process with semi-quantitative, ti/IFTA<25%, ti/IFTA 26-50%), ti/IFTA >50%. For the aforementioned reasons, IFTA assessment can suffer from suboptimal inter-and intra-variability among renal pathologists[1, 39–42]. Employing DL models to estimate IFTA offers a quantitative approach that has the potential to mitigate this variability. In this study, a senior nephropathology reviewed the external dataset comprising 24 WSIs and provided IFTA scores. Consistent with our expectations, the performance of the models tends to decline with increasing IFTA extent. This decline is reflected in the pathologists’ assessments of IFTA on renal biopsies, indicating that as the severity of IFTA increases, the models’ accuracy in predicting these scores correspondingly decreases. It is important to note that the majority of existing DL models have primarily been trained on biopsy specimens which do not have the vast array of pathological alterations, which does not encompass the full spectrum of renal pathology. Although these models have high accuracy and IoU in their test cohort, the models start to fail when tackling complex regions like fibrosis and inflammation. Our DL model has a good performance on the different grades of IFTA, including the easy group, medium group, and hard group in the external dataset. In the future, this underscores the need for more diverse and comprehensive datasets to enhance the accuracy and generalizability of DL models in the realm of renal pathology.

In AI segmentation algorithms, mIoU is a crucial metric that evaluates model performance by averaging the performance across all categories. The contribution of mIoU from each category to the collective outcome is substantial. If the model performs poorly in one or several categories, it can adversely impact the average class mIoU, despite good performance in other categories. In this study, on the macro-level, only acute tubular injury was found to correlate with a lower IoU across all 4 compartments. Therefore, in practical applications, optimizing the model for the acute tubular injury category could potentially enhance the average class segmentation performance. However, a deeper understanding of the histological parameters that negatively affect segmentation performance might identify case characteristics that can provide an active learning signal for future iterations of model improvement. These data suggest that in the setting of active learning, including new annotations with high degrees of total inflammation (ti-score) and interstitial fibrosis and tubular atrophy (IFTA-score) could negatively impact the IoU for interstitial segmentation or vice versa.

Furthermore, building on the basic anatomy segmentation model allows us to easily scale the model in complexity by adding classes that represent lesions. We have built on the concept of the human-AI-loop (HAIL)[43], utilizing the trained segmentation model to generate new annotations, practically creating instance segmentations via post-processing of multiclass semantic prediction masks. Such an approach could inspire the community to extend the segmentation models with increased complexity without much effort, similar to the recently open-sourced zero-shot general-purpose Segment Anything Model (SAM) for natural images[44].

In conclusion, we train an open-source segmentation model for the basic anatomical structure in kidney histology, and we demonstrate that Transformer-based models exhibit superior performance on generalizability with limited data, while UNet-based models can achieve comparable performance with the Transformer-based models if provided with sufficient data. Moreover, Swin-B is a better backbone encoder than the ResNet-18/50 encoders in renal anatomical segmentation. In the future, visualizing the decision-making process of pathologists and matching quantitative reports of specific renal biopsies with those from the archives, will enable lesion-based image retrieval and enhance diagnostic precision.

## Data availability statement

The source code and the model weights are available online https://github.com/amspath after request.

## Authors’contributions

He J and Kers J designed this study. Kers J provided critical intellectual input into the manuscript with revisions. He J, Long J and Li J annotated the WSIs, and Kers J reviewed and corrected the annotations. Florquin S, Naesens M, Priyanka Koshy, Nguyen TQ, Meziyerh S, De Vries APJ, De Boer OJ collected WSIs and performed histological scores according to the Banff lesion scoring system. He J, Valkema PA and Xiong Z trained and validated the DL-models. Valkema PA updated the annoation software-slidescape, He J, Valkema PA and Xiong Z interpreted the data and wrote the first draft of the manuscript. Verbeek FJ performed the data visualization. All authors contributed to data sorting, and read and approved the final version to be published.

## Funding Acknowledgement

This work was supported by grants from Chongqing Science and health joint medical research project (general program, 2024MSXM017) and the New Chongqing Talent Attraction Program Research Project (CQYC20230A04254).

---

**Figure S1.**
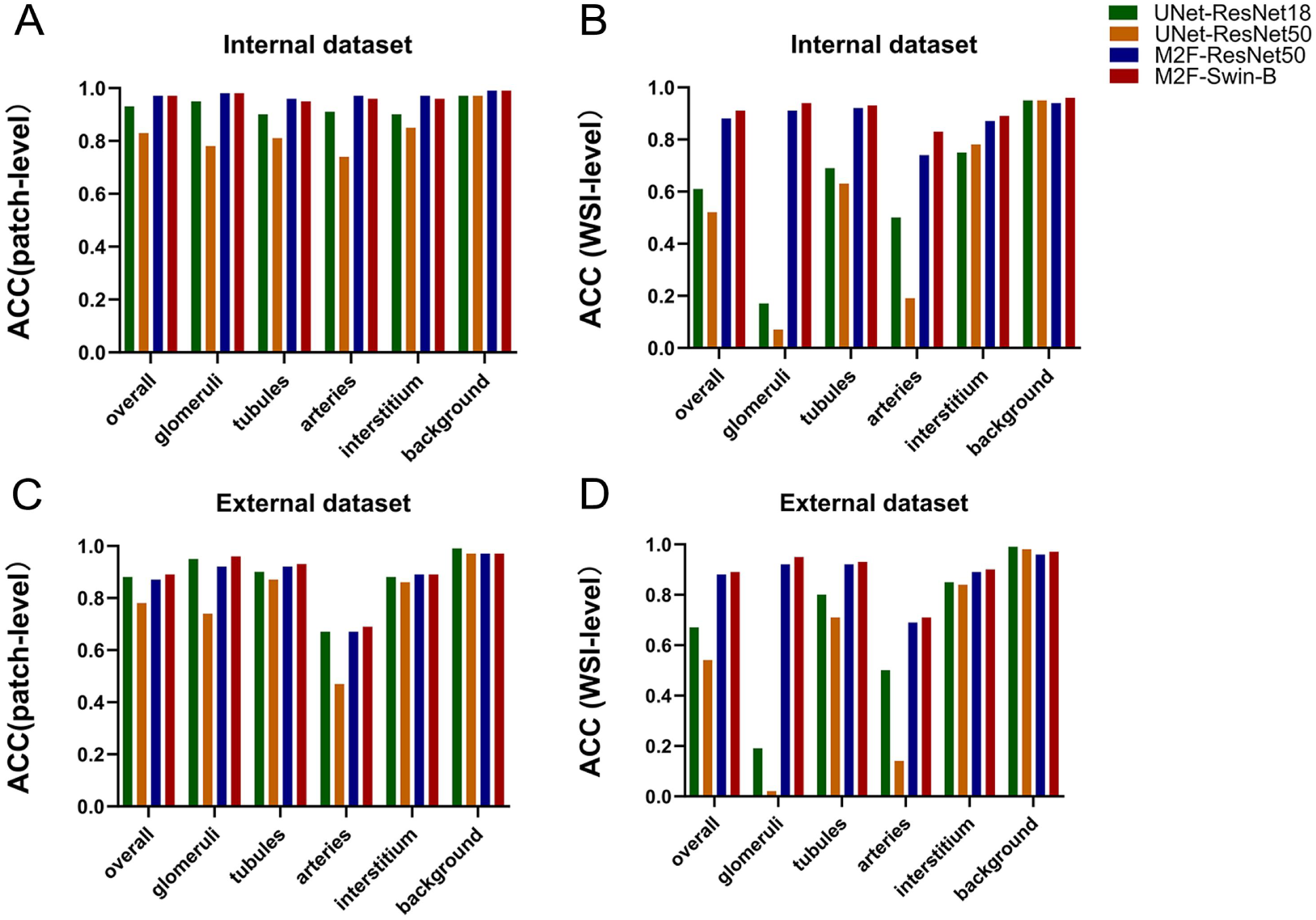
The performance of different DL models on the internal validation dataset with patch-level and WSI-level split. **A-B**. ACC of overall class and per-class of four models (UNet-ResNet18, UNet-ResNet50, M2F-ResNet50 and M2F-SwinB) in the internal dataset with the patch-level split (A) and WSI-level split (B). **C-D**. ACC of overall class and per-class of four models in the external dataset with the patch-level split (C) and WSI-level split (D). ACC: accuracy; M2F:Mask2Former.

**Figure S2.**
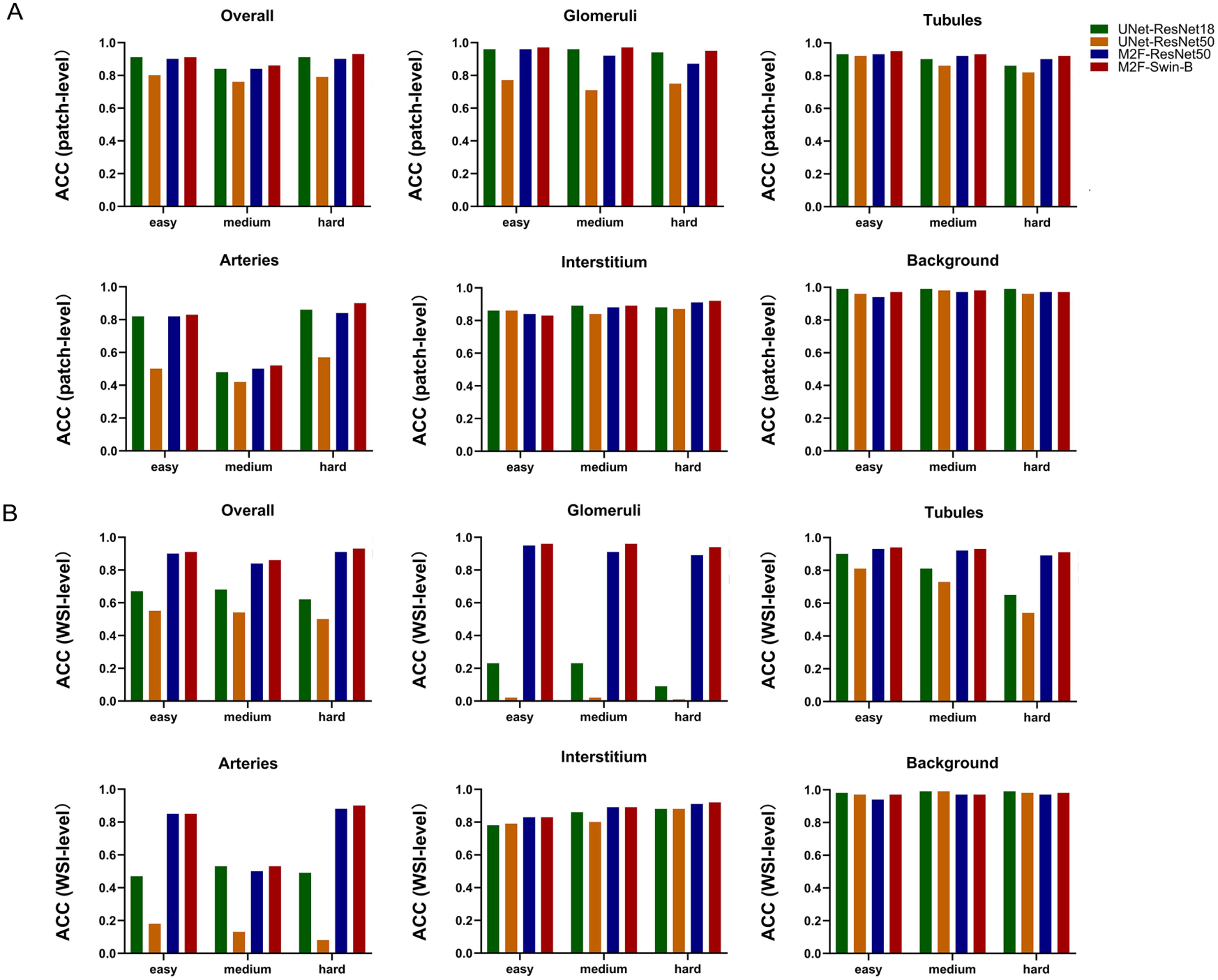
The performance of different models on the subgrouped external dataset with the patch-level split. **A**. ACC of overall class, glomeruli, tubules, arteries, interstitium, and background in the subgrouped external dataset (easy group, medium group, hard group) for different models with the patch-level split. **B**. ACC of overall class, glomeruli, tubules, arteries, interstitium, and background in the subgrouped external dataset (easy group, medium group, hard group) for different models with the WSI-level split. ACC: accuracy; M2F: Mask2Former.

**Figure S3.**
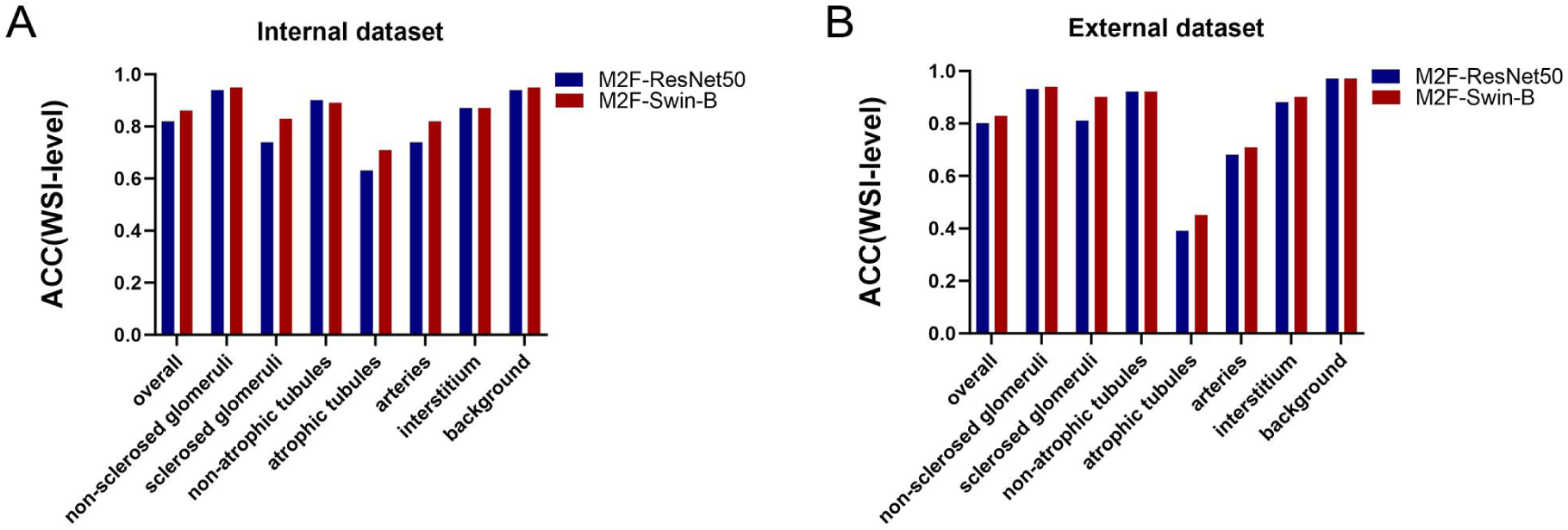
The performance of Mask2Former on segmenting the normal and abnormal kidney anatomical compartments. **A-B**. ACC of overall class, non-sclerosed glomeruli, sclerosed glomeruli, non-atrophic tubules, atrophic tubules, arteries, interstitium, background in two different Mask2Former models in the internal dataset (A) and external dataset (B) with the WSI-level split. ACC: accuracy; M2F: Mask2Former.

